# Effects of high-intensity interval training with or without resistance training on Alzheimer’s disease brain signatures in individuals with coronary artery disease: the Heart-Brain randomized controlled trial

**DOI:** 10.64898/2026.07.02.26357035

**Authors:** Javier Sanchez-Martinez, Patricio Solis-Urra, Angel Toval, Andrea Coca-Pulido, María Teresa Rodríguez Palacios, Lucía Sánchez-Aranda, Esmée A. Bakker, Isabel Martín-Fuentes, Javier Fernández-Ortega, Rosa María Alonso-Cuenca, Marcos Olvera-Rojas, Beatriz Fernandez-Gamez, Kirk I. Erickson, Eduardo Moreno-Escobar, Rocío García-Orta, Irene Esteban-Cornejo, Francisco B. Ortega

## Abstract

**Background:** Coronary artery disease (CAD) increases the risk of cognitive impairment, dementia, and brain structural alterations. This heart-brain connection suggests that exercise-induced cardiovascular responses may influence brain regions vulnerable to Alzheimer’s disease (AD), the most common cause of dementia. This study aimed to examine the effects of 12-weeks of either high-intensity interval training (HIIT) plus resistance training (RT) or HIIT alone on AD brain signatures in individuals with CAD, to explore moderating factors, and to assess associations between changes in AD brain signatures and cognition and physical fitness.

**Methods:** This secondary analysis of a single-site, three-arm, single-blinded randomized controlled trial included 105 individuals with CAD (50-75 years; 21% female) randomly allocated to HIIT+RT, HIIT, or usual care (UC) groups. T1- and diffusion-weighted magnetic resonance imaging were acquired before and after the 12-week intervention. Primary outcomes were thickness/volume and gray matter mean diffusivity (GMMD) signatures, derived from seven cortical regions and the hippocampus. Moderators included age, sex, education, and baseline AD brain signatures.

**Results:** For the thickness/volume signature, no between-group differences in changes were observed between HIIT+RT and UC (+0.13 standardized mean difference [SMD]; 95% CI, -0.07 to 0.33) or between HIIT and UC (-0.1 SMD; 95% CI, -0.3 to 0.1); however, a small but significant between-group difference in change was found between HIIT+RT and HIIT, in favor of HIIT+RT (+0.23 SMD; 95% CI, 0.03 to 0.42). For the GMMD signature, no significant between-group differences in changes were found between HIIT+RT and UC (+0.08 SMD; 95% CI, -0.18 to 0.34), HIIT and UC (+0.1 SMD; 95% CI, -0.16 to 0.36), or HIIT+RT and HIIT (-0.02 SMD; 95% CI, -0.28 to 0.23). No moderation effects were identified, and no associations were observed between changes in AD brain signatures and cognition or physical fitness.

**Conclusion:** A 12-week HIIT+RT intervention was more effective than HIIT alone in increasing the thickness/volume signature in individuals with CAD, yet no differences were observed compared to UC and the interventions did not affect the GMMD signature. These findings suggest that the intra-session inclusion of RT with HIIT may enhance AD-related brain macrostructure in individuals with CAD more than just HIIT training.

## 1. Introduction

Emerging evidence supports a heart-brain connection that may explain the impact of coronary artery disease (CAD) on brain health.^1^ CAD is associated with structural brain changes, including greater atrophy,^2,3^ and increased risk of cognitive impairment and all-cause dementia,^4,5^ including Alzheimer’s disease (AD).^6^ Given the elevated AD risk in individuals with CAD, identifying modifiable factors able to enhance brain health in this population is necessary. In this context, the AD brain signature represents a clinically relevant brain health outcome, reflecting a set of brain measures commonly implicated in AD. For instance, higher AD signature based on cortical thickness (macrostructure) is associated with lower dementia risk in cognitively unimpaired older adults.^7^ Furthermore, elevated gray matter mean diffusivity (GMMD) in AD-related regions, reflecting microstructural disruption, was associated with greater AD risk in cognitively unimpaired middle-aged adults.^8^ Therefore, identifying interventions that preserve brain integrity in individuals with CAD is crucial to prevent cognitive decline and reduce AD vulnerability.

Evidence links exercise to improved brain health,^9,10^ suggesting potential benefits in brain structure for individuals with CAD. A meta-analysis of 59 randomized controlled trials (RCTs) involving healthy and cognitively impaired populations reported increases in frontal and hippocampal volumes following exercise interventions.^9^ However, our recent meta-analysis indicated that studies examining the effects of exercise on brain structure in individuals with CAD are rare, despite improvements in mental health and quality of life. This gap highlights the need for further research focusing on brain outcomes.^11^ Recent American Heart Association (AHA) guidelines emphasize the combination of aerobic exercise and resistance training (RT) to optimize overall health.^12^ In individuals with CAD, a 6-month exercise-based cardiac rehabilitation program combining moderate-intensity continuous training (MICT) and RT increased gray matter volume in frontal, temporal, supplementary motor, and cerebellar regions; however, the lack of a control group precludes attributing these changes solely to exercise.^13^ Thus, despite the lack of RCTs, the impact of exercise on brain structure, including AD related regions, in CAD populations remains promising.

Nevertheless, the effects of exercise on AD-related brain regions may vary by protocol. For example, different exercise modalities have shown contrasting effects on the hippocampus in older adults. A 4-month MICT plus high-intensity interval training (HIIT) program tended to increase left hippocampal volume compared with low-intensity continuous training,^14^ whereas a 6-month HIIT program prevented reductions in the right hippocampal volume in comparison to MICT and low-intensity exercise.^15^ In contrast, a 5-year HIIT intervention reduced hippocampal volume compared with the control group, whereas no differences were observed between the MICT and control groups.^16^ Contrasting effects have also been reported for AD brain signatures. A 6-month RT program in cognitively unimpaired older adults reduced the thickness/volume signature in the amyloid-beta-positive subgroup but had no effect on the GMMD signature.^17^ Conversely, a 2-year multidomain lifestyle intervention including physical activity prevented decline in the cortical AD signature among older adults with elevated cardiovascular risk and low baseline cortical thickness.^18^ Therefore, evaluating the differential effects of exercise modalities is essential when exploring brain structure, including AD-related macro- and microstructural brain signatures, in individuals with CAD.

This study aimed to examine the effects of HIIT, with or without RT, on AD brain signatures, specifically thickness/volume and GMMD signatures, in individuals with CAD. Secondary aims included identifying possible moderating factors and assessing associations between changes in AD brain signatures and cognition and physical fitness. We hypothesized that (i) both exercise programs would increase thickness/volume signature and reduce GMMD signature, with the HIIT+RT potentially being superior; (ii) the effects of exercise interventions on AD brain signatures might be moderated by age, sex, education level, or baseline AD brain signature levels; and (iii) changes in these signatures would correlate with changes in cognition and physical fitness.

## 2. Materials and methods

### 2.1. Protocol registration, design, and participants

This study includes secondary analyses conducted within the framework of the Heart-Brain trial,^19^ registered on Clinicaltrials.gov (Identifier: NCT06214624; https://clinicaltrials.gov/study/NCT06214624) on December 22, 2023, including a prespecified statistical analysis plan. The Hearty-Brain trial is a single-site, three-arm, single-blind RCT. A total of 105 individuals with stable CAD were recruited from two public hospitals in Granada, Spain, from April 2022 to June 2024. Sample size was calculated for the primary outcome (cerebral blood flow) of the Heart-Brain trial.^19^ Further methodological details of the Heart-Brain trial have been previously described in the trial protocol.^19^ Standard operating procedures for the intervention and outcome assessments are publicly available on GitHub (https://github.com/Heart-Brain/Heart-Brain) and Zenodo (https://zenodo.org/records/17865709). Moreover, the effects of the intervention on the primary outcome (cerebral blood flow) and other secondary outcomes are available elsewhere.^20^ No deviations occurred in the study design, intervention, procedures, or the statistical analysis plan. The study followed the principles of the Declaration of Helsinki and was approved by the Research Ethics Board of the Andalusian Health Service (CEIM/CEI Provincial de Granada; #1776-N-21 on December 21st, 2021). All participants provided written informed consent once all study details were explained. The results are reported in accordance with the 2025 Consolidated Standards of Reporting Trials (CONSORT) statement (Supplementary Table 1).^21^

### 2.2. Eligibility criteria

Details of the inclusion and exclusion criteria were previously published in the study protocol.^19^ Briefly, eligibility criteria included: (i) individuals with chronic CAD and stable medical treatment, aged between 50-75 years, (ii) with left ventricular ejection fraction ≥ 45%, (iii) physically inactive (not meeting the World Health Organization’s (WHO) weekly physical activity recommendations,^22^ and not participating in a planned and structured exercise program at least 3 days per week for the last 3 months), and (iv) ≥ 26 points in the Spanish version of the modified Telephone Interview of Cognitive Status (STICS-m),^23^ to exclude major cognitive impairment.

### 2.3. Randomization

Participants were randomized by a blinded external researcher using a 1:1:1 allocation ratio and stratified by age (<65 or ≥65 years) and sex (female or male) to one of three groups: HIIT+RT, HIIT and usual care (UC). Further details of the randomization procedure and blinding are provided in the trial protocol.^19^

### 2.4 Interventions

A detailed description of the Heart-Brain exercise interventions, following the Consensus on Exercise Reporting Template (CERT), is available elsewhere.^24^ Briefly, the HIIT+RT and HIIT programs were designed to be isotemporal (45 min) and isocaloric (∼775 MET-min/week), aligning with the 2020 WHO physical activity guidelines (≥600 MET-min/week). Moreover, both exercise programs were designed in accordance with guidelines for the delivery and monitoring of HIIT in clinical populations.^25^ Participants assigned to either exercise group attended 45-min supervised training sessions at the Sport and Health University Research Institute (iMUDS), University of Granada (Spain), three times per week for 12 weeks. Exercise sessions were led by accredited expert trainers with a BSc degree in sports sciences, using a 1:1 trainer-to-participant ratio.

#### 2.4.1. HIIT+RT intervention

Supervised sessions consisted of: i) a 5-min warm-up; ii) three 4-min HIIT bouts on a treadmill (85-95% HRmax) with 3-min recovery periods between bouts (70% HRmax); iii) a 4-min cool-down with a 1-min 20-s transition; and iv) two sets of an 8-exercise RT circuit including a combination of upper and lower limb exercises using elastic bands and body weight (20 s per exercise, 40 s rest between exercises, and 1-min rest between sets).

#### 2.4.2. HIIT intervention

Supervised sessions consisted of: i) a 10-min warm-up; ii) four 4-min HIIT bouts on a treadmill (85-95% HRmax) with 3-min recovery period between bouts (70% HRmax); and iii) a 10-min cool-down.

#### 2.4.3. UC group

Patients received usual medical care, including periodic medical visits during which clinicians could provide lifestyle and clinical advice and adjust medications as needed. After completing all assessments at the end of the 12-week period, participants were offered enrollment in the supervised HIIT program.

### 2.5. Image acquisition and preprocessing

Magnetic resonance imaging (MRI) was performed on a 3T scanner (Siemens Magnetom PRISMA Fit) equipped with a 64-channel head coil at the Mind, Brain and Behavior Research Centre (CIMCYC), University of Granada. T1-weighted magnetization-prepared rapid acquisition gradient echo (MPRAGE) sequences were acquired for structural brain scans (parameters: sagittal, 0.8 mm isotropic resolution, echo/inversion/repetition times = 2.31/1060/2400 ms, field of view = 256 mm, 224 slices; acquisition time: 6 min). Diffusion-weighted images were acquired using a diffusion sequence (parameters: Resolution: 2 × 2 × 2 mm, TE/TR = 95.6/2800 ms, multiband factor = 4, b-values of 1500, 3000 s/mm^2^, 64 gradient directions; acquisition time: 8 min). For structural and diffusion preprocessing, we replicated an approach previously applied in another sample.^17^ For details on preprocessing and quality control (QC), see Supplementary Methods.

### 2.6. Outcome measurements

#### 2.6.1. Alzheimer’s disease brain signatures

Thickness/volume and GMMD signatures were computed following Williams et al. (2021) methodology.^8^ We have previously applied this approach in a sample of cognitively unimpaired older adults.^17^ We selected this approach because it considers both macrostructural and microstructural AD signatures, in contrast to other approaches focused solely on macrostructural AD signatures. The thickness/volume signature comprises a weighted average of cortical thickness in seven ROIs (entorhinal cortex, middle temporal gyrus, bank of superior temporal sulcus, superior temporal gyrus, isthmus cingulate, lateral orbitofrontal cortex, and medial orbitofrontal cortex), along with hippocampal volume (Figure 1). The weights assigned to each ROI for the left and right hemispheres are detailed in Williams et al. (2021). Structural and diffusion data for each ROI were adjusted for age, and hippocampal volume was further regressed for estimated intracranial volume to account for differences in head size. Standardized residuals for each ROI were computed separately at each time point (baseline and post assessment), then weighted and summed to calculate the thickness/volume signature scores. The same weights used for structural data were applied to the GMMD values of each ROI. Further details of the MRI processing are available in Supplementary Material.

**Figure 1.**
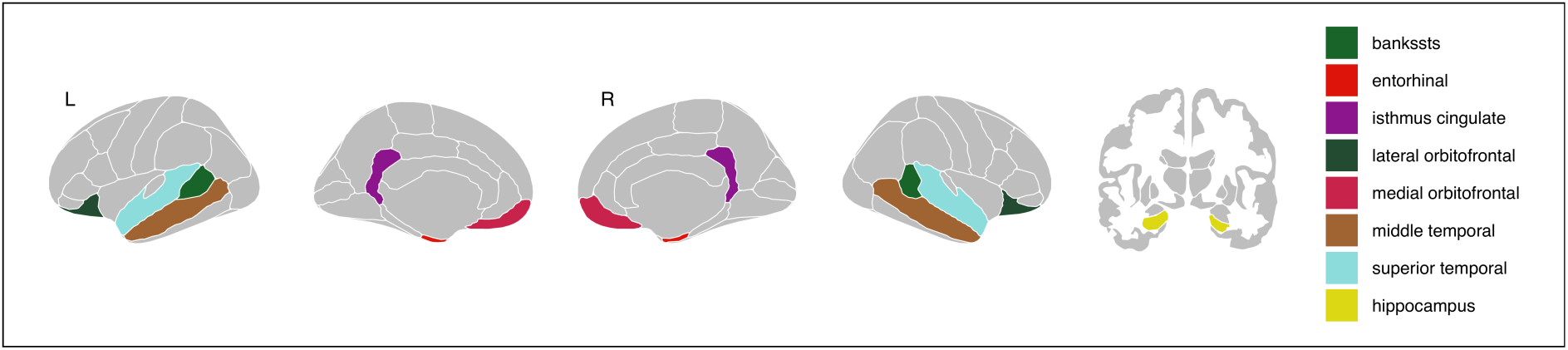
Brain regions used to compute Alzheimer’s disease brain signatures. Abbreviation: bankssts, bank of superior temporal sulcus.

#### 2.6.2. Cognitive outcomes

At baseline and after the intervention, participants completed a comprehensive neuropsychological evaluation that measured global cognition assessed by the Montreal Cognitive Assessment (MoCA)^26^ and several different cognitive domains (Supplementary Table 2). Details of the cognitive tests have been previously described.^19^ Composite scores for each cognitive domain were computed using multiple cognitive indicators.^27^ Z-scores adjusted to baseline were calculated for episodic memory, executive function/attentional control, processing speed, and working memory, following the approach used in previous analyses of this sample.^27^

#### 2.6.3. Physical fitness

At baseline and after the intervention, participants performed a standardized ramp incremental cardiopulmonary exercise test on a treadmill (h/p/cosmos, Nussdorf, Germany), according to the American College of Sports Medicine guidelines.^28^ Respiratory gases were measured using a dilution flow system (Omnical, Maastricht Instruments, Maastricht, the Netherlands). Relative VO_2_peak (mL/kg/min) was used as the indicator of cardiorespiratory fitness. Additional details are provided elsewhere.^27^ Lower-body muscular endurance was assessed with the 30-s Chair Stand Test (number of repetitions) following the Senior Fitness Test battery procedures.^29^

### 2.7. Statistical analysis

#### 2.7.1. Main analysis

All analyses were performed using R (v4.5.1). Constrained longitudinal linear mixed models (baseline adjusted) were fitted using the “*lmerTest*”^30^ and “*lme4*”^31^ packages with restricted maximum likelihood estimation to assess the intervention effects on thickness/volume and GMMD signatures. Models included time as fixed effect, and group-by-time interaction with random intercepts and unequal variance across time and group. Statistical significance was set at *p* <0.05 without adjustments for multiple comparisons. Estimated marginal means, within-group changes, and between-group differences were extracted from the model using the “*emmeans”* package.^32^. An intention-to-treat approach was used as the primary analysis, while per-protocol analysis (≥ 70% session attendance, defined as the proportion of sessions completed out those offered) as secondary/exploratory, as we specified in our protocol paper.^19^ As pre-specified in our statistical analysis plan, missing data were assumed to be missing at random and were handled within the linear mixed models. Baseline and post-intervention z-scores were computed using the baseline mean and standard deviation (SD) of the sample. Changes in z-scores for each intervention were then computed, representing standardized mean differences (SMDs) that quantify how post-intervention values deviate from baseline. Main intervention effects were evaluated by comparing between-group differences in z-score change from baseline to 12 weeks.

#### 2.7.2. Sensitivity analysis

For the thickness/volume signature, a sensitivity analysis was conducted by excluding images with parcellation issues in the ROIs used to compute the signature, as determined by visual quality control following the ENIGMA Consortium Cortical QC Protocol 2.0 (https://enigma.ini.usc.edu). In addition, we examined the effects of the interventions on AD brain signatures derived from cortical thickness and volumes, incorporating six alternative methodologies that included additional brain regions (see Supplementary Fig. 1).^33–38^ Additionally, we tested intervention effects after excluding participants with only pre-intervention T1 images (n=15) to account for potential variability arising from the use of only the cross-sectional FreeSurfer recon-all pipeline instead of the longitudinal pipeline available for subjects with both pre- and post-intervention images (see Supplementary Methods).

For the GMMD signature, sensitivity analyses excluded low-quality images identified through visual QC, automated QC, and those with incorrect acquisition parameters.

#### 2.7.3. Moderator effects

Moderation was examined by treating age, sex, education level, and outcome baseline level as interaction terms. Age was categorized as younger (<65 years) and older (≥65 years). Sex was categorized as male and female. Education level was classified as Primary/Secondary and University. Thickness/volume signature and GMMD signature levels were categorized as high or low based on the baseline median. Significance was set at *p* <0.05.

#### 2.7.4. Associations of changes

Unadjusted linear regression models were used to examine the associations between z-score changes in thickness/volume and GMMD signatures (predictors, analyzed separately) and changes in cognition (global cognition and cognitive domains) and changes in physical fitness outcomes (dependent variables). Statistical significance was set at *p* <0.05.

## 3. Results

Table 1 presents the baseline characteristics of the sample. The mean age of the participants was 62.1 years; most were male (79%), had primary or secondary education (60%), and had a mean BMI in the overweight range. Participant flow, from enrollment, allocation and analysis is summarized in Supplementary Fig. 2, while inclusion and exclusion based on brain image quality are detailed in Supplementary Results and Supplementary Table 3. Details about adherence, intensity compliance, enjoyment and affect response to the exercise programs have been previously described,^24^ as well as the adverse effects.^20^

**Table 1.** Baseline characteristics of the sample.

| Characteristics | n | Overall | HIIT+RT<br>(n = 37) | HIIT<br>(n = 35) | UC<br>(n = 33) |
| --- | --- | --- | --- | --- | --- |
| Age | 105 | 62.1 (6.6) | 62.0 (7.0) | 61.9 (6.6) | 62.5 (6.3) |
| Sex | 105 |  |  |  |  |
| Male |  | 83 (79%) | 29 (78%) | 27 (77%) | 27 (82%) |
| Female |  | 22 (21%) | 8 (22%) | 8 (23%) | 6 (18%) |
| Education level | 105 |  |  |  |  |
| Primary/Secondary school |  | 63 (60%) | 23 (62%) | 23 (66%) | 17 (52%) |
| University |  | 42 (40%) | 14 (38%) | 12 (34%) | 16 (48%) |
| BMI | 105 | 29.1 (3.9) | 29.0 (3.6) | 29.1 (4.3) | 29.2 (3.7) |
| APOE carrier | 105 |  |  |  |  |
| Non-carrier |  | 86 (82%) | 29 (78%) | 30 (86%) | 27 (82%) |
| Carrier |  | 19 (18%) | 8 (22%) | 5 (14%) | 6 (18%) |
| Moderate-vigorous PA (min/wk) | 104 | 320.8 (193.8, 452.3) | 326.5 (200.2, 437.6) | 373.5 (237.3, 477) | 298.8 (156.9, 362.6) |
| Self-reported muscle-strengthening (sessions/wk) | 98 | 0 (0, 3) | 0 (0, 3) | 0 (0, 4) | 0 (0, 3) |
| Telephone Interview of Cognitive Status | 105 | 35 (33, 36) | 35 (33, 36) | 35 (33, 36) | 35 (33, 36) |
| Cognition |  |  |  |  |  |
| Global cognition (MoCA) | 105 | 24.8 (3.3) | 24.6 (3.6) | 24.5 (3.4) | 25.3 (2.6) |
| Episodic memory (z-score) | 105 | 0.00 (1.00) | 0.11 (1.01) | -0.19 (1.04) | 0.08 (0.95) |
| EF & attentional control (z-score) | 105 | 0.00 (1.00) | -0.01 (0.96) | -0.12 (1.26) | 0.13 (0.71) |
| Processing speed (z-score) | 105 | 0.00 (1.00) | -0.05 (1.02) | -0.06 (1.17) | 0.12 (0.77) |
| Working memory (z-score) | 103 | 0.00 (1.00) | -0.08 (0.98) | -0.02 (1.15) | 0.12 (0.86) |
| Physical fitness |  |  |  |  |  |
| VO<sub>2peak</sub> (mL/kg/min) | 103 | 25.2 (5.0) | 25.2 (4.9) | 26.0 (5.2) | 24.5 (5.0) |
| 30-s Chair Stand Test (repetitions) | 105 | 16 (14, 19) | 16 (13, 18) | 16 (15, 19) | 15 (14, 19) |
| AD brain signatures (z-score) |  |  |  |  |  |
| Thickness/volume signature | 96 | 0.00 (1.00) | -0.29 (0.94) | 0.26 (1.09) | 0.03 (0.91) |
| GMMD signature | 94 | 0.00 (1.00) | 0.14 (1.16) | -0.17 (0.94) | 0.04 (0.88) |

Data are shown as mean (standard deviation), n (%), or median (Q1, Q3). Due to missing or invalid data, the sample size (n) varied across outcomes as follows: moderate-vigorous PA: HIIT+RT = 36, HIIT = 35, UC = 33; self-reported muscle-strengthening: HIIT+RT = 33, HIIT = 35, UC = 30; working memory: HIIT+RT = 37, HIIT = 34, UC = 32; VO<sub>2peak</sub>: HIIT+RT = 37, HIIT = 33, UC = 33; thickness/volume signature: HIIT+RT = 33, HIIT = 33, UC = 30; GMMD signature: HIIT+RT = 31, HIIT = 33, UC = 30. Abbreviations: AD, Alzheimer's disease; APOE4, apolipoprotein E ε4; BMI, body mass index; EF, executive function; GMMD, gray matter mean diffusivity; HIIT, high-intensity interval training group; HIIT+RT, high-intensity interval training plus resistance training group; min, minutes; MoCA, Montreal cognitive assessment; PA, physical activity; UC, usual care group; VO<sub>2peak</sub>, peak oxygen uptake; wk, week.

### 3.1. Effects on thickness/volume signature

In the primary intention-to-treat analysis, no significant between-group differences in changes in the thickness/volume signature were observed between HIIT+RT and UC (SMD = +0.13 [95% CI, -0.07 to 0.33]; p = 0.20) or between HIIT and UC (SMD = -0.1 [95% CI, -0.3 to 0.1]; p = 0.35) (Figure 2A). However, a small but significant between-group difference in change was observed between HIIT+RT and HIIT, favoring HIIT+RT (SMD = +0.23 [95% CI, 0.03 to 0.42]; p = 0.02). In the per-protocol analysis, two participants from the HIIT+RT group and two from the HIIT group were excluded, and the results were similar, with a trend toward a greater increase in HIIT+RT versus HIIT (SMD = +0.19 [95% CI, -0.01 to 0.39]; p = 0.06) (Supplementary Table 4).

**Figure 2.**
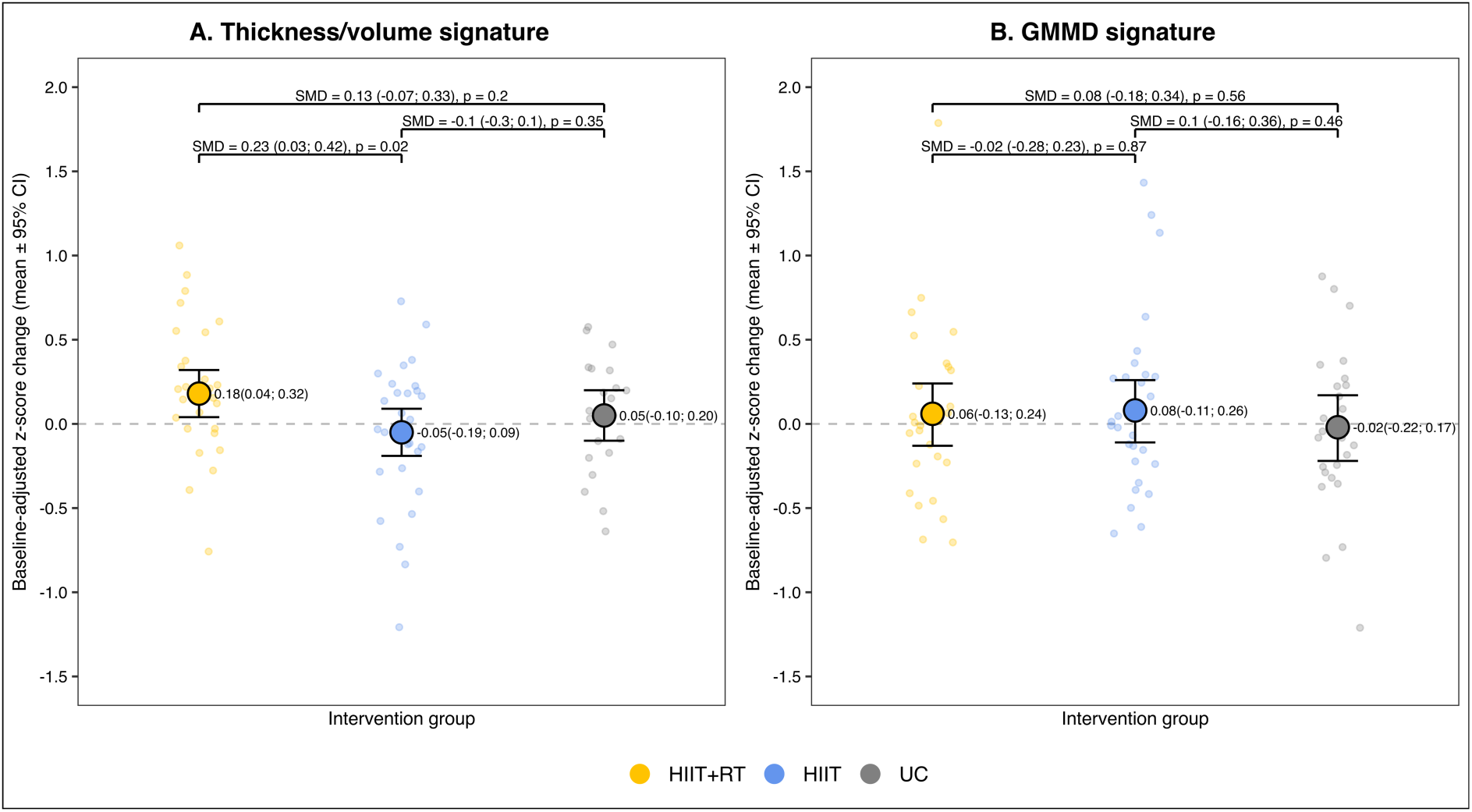
Effects of 12-week exercise interventions on Alzheimer’s disease brain signatures in individuals with coronary artery disease. Baseline-adjusted z-score change (mean ± 95% confidence intervals) for (A) the thickness/volume signature and (B) the gray matter mean diffusivity signature. Individual dots represent unadjusted z-score changes. Higher thickness/volume signature is associated with lower dementia risk; therefore, increments could be interpreted as protective. Elevated gray matter mean diffusivity in AD-related regions, reflecting microstructural disruption, is associated with greater Alzheimer’s disease risk, therefore, increments could be interpreted as a risk increase. Abbreviations: CI, confidence interval; GMMD, gray matter mean diffusivity; HIIT, high-intensity interval training group; HIIT+RT, high-intensity interval training plus resistance training group; SMD, standardized mean difference; UC, usual care group.

Likewise, sensitivity analyses were consistent with the main intention-to-treat findings, showing a significant increment in HIIT+RT versus HIIT (SMD = +0.27 [95% CI, 0.07 to 0.47]; p = 0.01) when low-quality images were excluded (Supplementary Table 5), as well as when participants with only pre-intervention images were excluded (SMD = +0.25 [95% CI, 0.05 to 0.45]; p = 0.02) (Supplementary Table 6). Analyses using six alternative methods to compute the AD brain signature (based on cortical thickness or volume) were consistent with the main findings. Across all six methods, HIIT+RT showed significant increases compared with HIIT (SMDs = +0.21 to +0.28; all p ≤ 0.03). Notably, two methods additionally showed significant increases of HIIT+RT versus UC (SMDs = +0.15 to +0.22; all p < 0.04) (Supplementary Table 7). Age, sex, educational level, and baseline outcome levels did not moderate the effects on the thickness/volume signature (Figure 3A).

**Figure 3.**
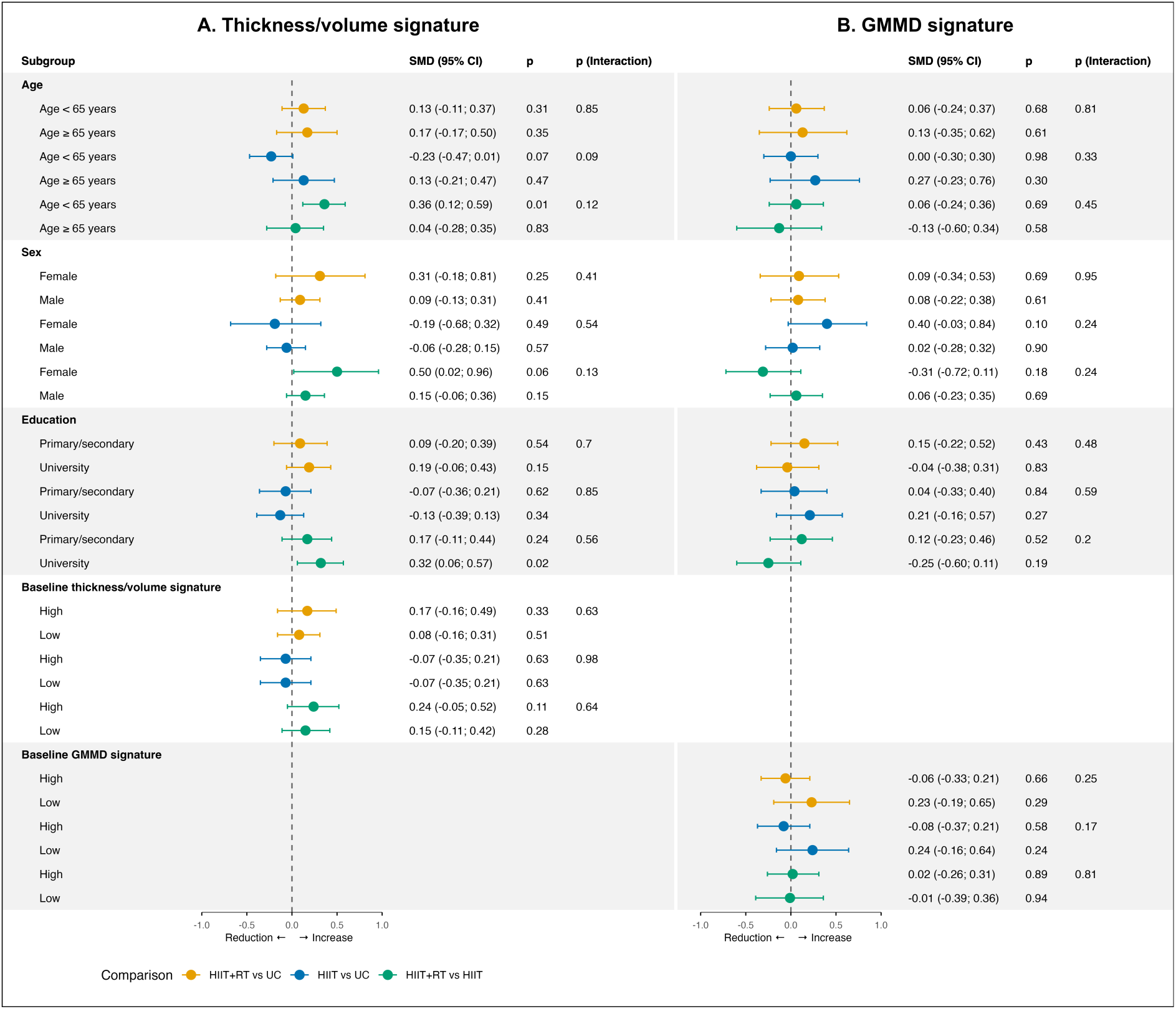
Moderator analyses of the effect of 12-week exercise interventions on Alzheimer’s disease brain signatures in patients with coronary artery disease. P (Interaction) indicates the interaction of the subgroup categories in the same group comparison; for example, the interaction of age categories for the comparison between HIIT+RT vs Control. Abbreviations: GMMD, gray matter mean diffusivity; HIIT, high-intensity interval training group; HIIT+RT, high-intensity interval training plus resistance training group; SMD, standardized mean difference; UC, usual care group.

### 3.2. Effects on GMMD signature

For the GMMD signature, no significant between-group differences in changes were observed in the intention-to-treat analysis between HIIT+RT and UC (SMD = +0.08 [95%CI, -0.18 to 0.34]; p = 0.56), HIIT and UC (SMD = +0.1 [95%CI, -0.16 to 0.36]; p = 0.46), or HIIT+RT and HIIT (SMD = -0.02 [95%CI, -0.28 to 0.23]; p = 0.87) (Figure 2B). In the per-protocol analysis, two participants from the HIIT+RT group and two from the HIIT group were excluded, and results remained consistent with the intention-to-treat findings (Supplementary Table 4). Sensitivity analyses excluding low-quality images (Supplementary Table 5) or participants with only pre-intervention images (Supplementary Table 6) also yielded consistent results. No moderator effects on the GMMD signature were identified (Figure 3B).

### 3.3. Associations between changes in AD brain signatures and cognition and physical fitness

No associations were found between changes in physical fitness outcomes and changes in the thickness/volume signature or the GMMD signature (Figure 4A). However, a trend toward a positive association was observed between changes in the thickness/volume signature and lower-body muscular endurance (p = 0.07).

**Figure 4.**
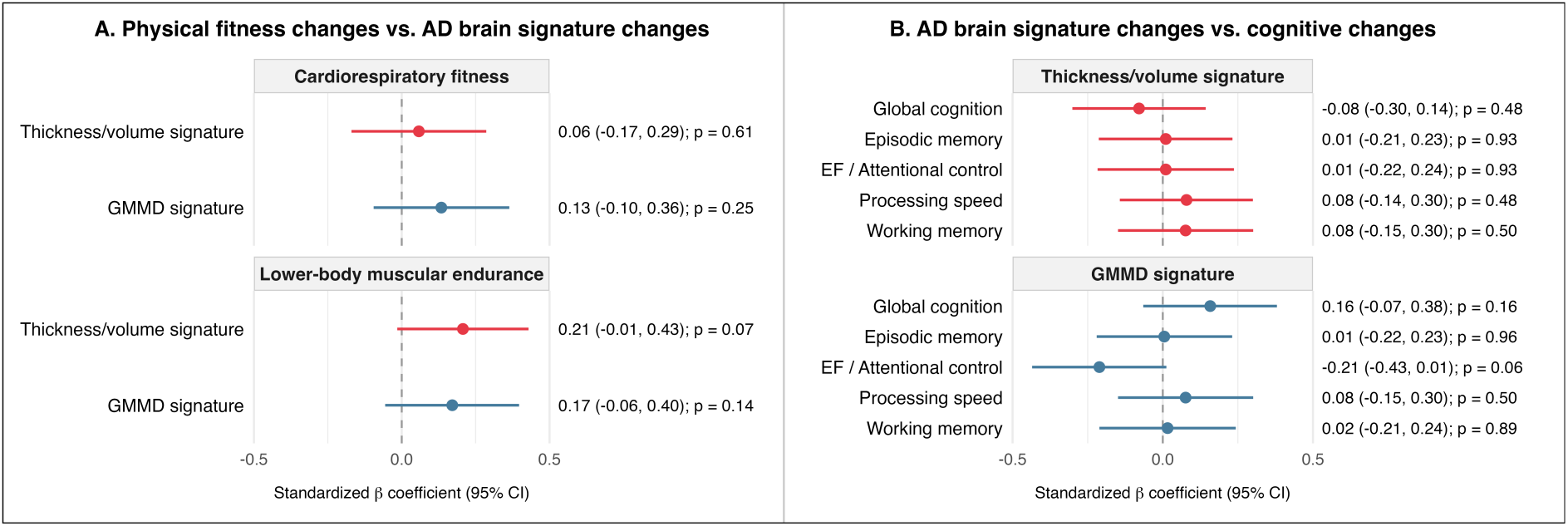
Associations between changes. (A) Associations between changes in physical fitness and changes in the Alzheimer’s disease brain signature changes. (B) Associations between changes in the Alzheimer’s disease brain signature and changes in cognition. Abbreviations: AD, Alzheimer’s disease; CI, confidence interval; GMMD, gray matter mean diffusivity.

No associations were found between changes in the thickness/volume signature or the GMMD signature and changes in cognitive outcomes (Figure 4B). However, a trend for a negative association was observed between changes in GMMD signature and executive function/attentional control (p = 0.06).

## 4. Discussion

This study examined the effects of 12 weeks of HIIT, combined with or without RT, on AD brain signatures in individuals with CAD, and explored potential moderators and associations with cognition and fitness. Regarding the thickness/volume signature, no between-group differences in the changes were observed between HIIT+RT or HIIT versus UC. However, and consistent with our hypothesis, HIIT+RT was superior to HIIT in increasing the thickness/volume signature. This finding showed the same direction in a per-protocol analysis. In contrast, no between-group differences were observed for the changes in the GMMD signature. In addition, age, sex, education level, and baseline outcome levels did not moderate the effects on AD brain signatures. Finally, changes in AD brain signatures were not associated with either cognition or physical fitness.

Few studies have examined how exercise interventions influence AD brain signatures, and existing results are heterogeneous. Regarding the thickness/volume signature, our findings are consistent with a 2-year multidomain lifestyle intervention, including muscle strength training and aerobic exercise, that prevented the decline in the cortical AD signature among older adults with elevated cardiovascular risk and lower baseline cortical thickness.^18^ In contrast, in our previous trial, a 6-month RT intervention in cognitively unimpaired older adults reduced the thickness/volume signature, although only among amyloid-beta-positive participants.^17^ Notably, a 3-month exercise intervention in older adults, including moderate-intensity exercises, RT and dual-task exercises, increased thickness and volume in prefrontal regions associated with AD and prevented hippocampal atrophy.^39^ These findings suggest that even short-term exercise interventions may induce structural brain changes. For the GMMD signature, our results are consistent with those of the aforementioned 6-month RT intervention, which showed no effect.^17^ This suggests that our exercise interventions did not influence AD-related brain microstructure, but it remains unknown whether longer interventions or other exercise modalities may affect these regions. Moreover, this study was not powered to detect small effect sizes; therefore, adequately powered studies evaluating different exercise modalities are needed to determine their effects on brain microstructure in AD-related regions. Overall, exercise interventions combining aerobic and RT may prevent decline or increase thickness and volume in AD-related regions among individuals with elevated cardiovascular risk, including those with CAD.

The increase in the thickness/volume signature after 12 weeks of HIIT+RT, compared with HIIT alone, may reflect a crucial benefit of including RT. Consistently, only the HIIT+RT group improved lower-body muscular resistance,^20^ and we observed a positive association between the thickness/volume signature and this fitness outcome, showing a trend toward statistical significance (p = 0.07). Importantly, our findings support the inclusion of RT to exercise programs, the benefits of which for optimizing health have been emphasized by a recent AHA scientific statement.^12^

Potential mechanisms underlying the impact of adding RT to promote brain plasticity include activation of the muscle-brain axis, vascular adaptations, and reduced inflammation. Skeletal muscle contractions stimulate the release of myokines into the bloodstream, including irisin, insulin-like growth factor 1 (IGF-1), and brain-derived neurotrophic factor (BDNF), which may promote brain plasticity.^40,41^ Studies have reported increases in IGF-1 and neurotrophin-4 levels following RT,^42–44^ as well as elevations in neurotrophin-3 levels and BDNF after RT or combined training programs including RT,^44–46^ highlighting the potential role of RT in stimulating the muscle-brain axis. Vascular adaptations may also contribute to these effects. Arterial stiffness has been associated with lower AD signature volumes,^47^ and both 6- and 12-week HIIT+RT interventions have been shown to reduce arterial stiffness in hypertensive adults and in individuals with chronic heart failure.^48,49^ Interestingly, a resistance-type HIIT session was more effective in reducing arterial stiffness than an aerobic-type (cycling) HIIT session,^50^ suggesting that RT may enhance the vascular benefits of HIIT. Another potential mechanism involves inflammation. Prospective evidence indicates that greater systemic inflammation during midlife is associated with smaller hippocampal and AD signature volumes in later life.^51^ RT has been shown to reduce circulating pro-inflammatory biomarkers, including tumor necrosis factor alpha, interleukin-6, interleukin-1β, and C-reactive protein, while increasing anti-inflammatory interleukin-10 levels in older adults.^44^ Therefore, the anti-inflammatory effects of RT may have contributed to the thickness/volume signature gains observed following HIIT+RT. Collectively, these mechanisms may influence cellular remodeling within the underlying architecture of AD-related brain regions and thereby contribute to changes in the thickness/volume signature observed after the HIIT+RT intervention. Studies examining these mechanisms in individuals with CAD remain limited; therefore, further research is needed to clarify the biological pathways underlying the effects of HIIT+RT on the thickness/volume signature.

Notably, although both interventions met the WHO aerobic activity guidelines (≥600 MET-min/week), only HIIT+RT fulfilled the additional recommendation of at least two weekly RT sessions. Despite increases in cardiorespiratory fitness after 12 weeks in both the HIIT+RT and HIIT groups, only the HIIT+RT group improved lower-body muscular endurance, as measured by the 30-s chair stand test,^20^ supporting the relevance of including RT in exercise interventions. A recent meta-analysis reported that interventions meeting WHO physical activity guidelines increased brain volume compared with those that did not;^9^ however, their definition of guideline adherence did not include RT. Therefore, the main contribution of this study to the existing literature is that adding RT to HIIT, thereby meeting full WHO guidelines and AHA recommendations, appears to be more effective than HIIT alone in increasing the thickness/volume signature in individuals with CAD. This finding was consistent and robust across six different methods used to compute the macrostructural AD signature. Although no between-group differences in changes were observed between HIIT+RT and UC for the thickness/volume signature, increases were identified in the HIIT+RT group compared with both HIIT and UC groups when two additional methods of deriving the macrostructural AD brain signature (based on cortical thickness or volume) were applied. This supports an effect of combined HIIT+RT and highlights the relevance of the methodological approach used to compute the AD brain signature, as it may influence the interpretation of the results.

AD brain signatures have been proposed as predictors of dementia risk, with greater AD-signature cortical thickness and lower GMMD signature associated with lower risk.^7,8^ After the 3-month intervention, we found no significant associations between changes in AD brain signatures and cognition in individuals with CAD. Although the current study shows HIIT+RT increased the thickness/volume signature, this did not translate into cognitive benefits.^20^ Consistent with these findings, a 2-year multidomain lifestyle intervention in older adults prevented the decline in the AD cortical signature without affecting cognition.^18^ In contrast, our previous study in cognitively unimpaired older adults reported that reductions in thickness/volume signature were associated with improvements in executive function.^17^ Regarding GMMD signature, we observed a trend toward a negative association with executive function/attentional control, suggesting that decreases in GMMD signature may be linked to cognitive benefits. This aligns with our prior study showing that reductions in GMMD signature were associated with attentional/inhibitory control improvements.^17^ Collectively, these findings suggest that the interpretation of AD brain signature changes may depend on age, disease stage, and signature modality, potentially reflecting non-linear trajectories across the AD continuum.^8^ Future studies combining AD brain signatures with other AD-related biomarkers are needed to clarify the biological and clinical significance of short-term intervention-induced changes.

## 5. Strengths and limitations

On the one hand, the study has several notable strengths. A key strength of this study was the use of both structural and diffusion brain imaging to assess exercise effects on novel AD brain signatures. The robustness of our findings across multiple sensitivity analyses and the usage of six additional methods to compute the macrostructural AD brain signature strengthens the credibility of the conclusions. Furthermore, the assessment and intervention protocols of the Heart-Brain trial are publicly available, as well as the structural and diffusion pipelines to enhance reproducibility. The exercise protocols themself are reproducible, and applicable to individuals with CAD. Additionally, the exercise interventions were randomly assigned and performed in a supervised setting. On the other hand, several limitations need to be acknowledged. The study focused on individuals with CAD, so the results may not be generalizable to other populations. The lack of follow-up prevented assessment of potential residual effects. Larger-scale interventions with extended follow-up are needed to confirm the effects of exercise on AD-related macro- and microstructural brain signatures. As MRI-derived outcomes were secondary outcomes of the trial, no power or sample calculations were performed. Finally, there were few participants in some subgroup categories, limiting interpretation of moderation results.

## 6. Practical/clinical implications

Our findings highlight the potential of a combined HIIT plus RT intervention to increase the thickness/volume signature compared to HIIT training alone, which may reflect neuroprotective effects in individuals with CAD who are at higher risk of dementia. The differential effects of exercise protocols could help guide the selection of optimal interventions for this population. Because the benefits were independent of age, sex, educational level, and baseline AD brain signature levels, this combined intervention could be applied broadly to support brain health. Moreover, our model is time-efficient, meeting aerobic and RT guidelines in just three 45-minute sessions, and demonstrates high attendance and compliance, no major adverse events, and safety,^20^ supporting its integration into cardiac rehabilitation programs. Further studies should investigate residual effects after exercise interventions, evaluate whether other exercise modes or longer intervention periods enhance the impact on AD brain signatures, and assess their clinical relevance.

## 7. Conclusion

A 12-week HIIT+RT intervention was more effective than HIIT alone in increasing the thickness/volume signature in individuals with CAD, yet no differences were observed compared to UC and the interventions did not affect the GMMD signature. These findings suggest that the intra-session inclusion of RT together with HIIT may enhance AD-related brain macrostructure in individuals with CAD compared to HIIT alone, potentially reflecting neuroprotective effects and possible implications for dementia risk, which should be confirmed in future long-term studies.

## Data Availability

Metadata (e.g. study protocols, data dictionaries, data processing scripts, statistical analyses plan) of the project will be open access available when the final script is ready after the peer-review process. All metadata will be hosted on a GitHub repository for version control and ease of reuse (https://github.com/Heart-Brain/Heart-Brain) and archived on Zenodo (https://doi.org/10.5281/zenodo.17865709) to ensure long-term preservation and to provide digital object identifiers (DOIs). The individual patient data of the Heart-Brain trial will not be made open access, due to privacy concerns and violation of the General Data Protection Regulation (due to the low number of cases in certain specific characteristics). Individual data will be available under restricted access following the "as open as possible, as closed as necessary" principle. The data files will include pseudonymized identification codes and only contain participants who provided informed consent for data sharing. The procedure for data sharing can be requested via the PI of the study (Prof. Ortega). Following this procedure, this study fully complies with the Open Science principles including FAIR data management, reproducibility, and inclusive, collaborative practices.

https://zenodo.org/records/17865709

## Authors’ contributions

JSM contributed to the design/conception, data collection, image processing, data analysis, data interpretation, manuscript preparation, and revision; PSU and IEC contributed to the design/conception, image processing, data analysis, data interpretation, manuscript preparation, and revision; FBO contributed to the design/conception, data analysis, data interpretation, manuscript preparation, and revision; LSA and JFO were involved in the intervention application, data collection, and manuscript revision; AT contributed to project management and manuscript revision; ACP, MOR, and BFG contributed to data collection and manuscript revision; EAB contributed to statistical analysis and manuscript revision; MTRP, IMF, RMAC, KIE, EME and RGO contributed to manuscript revision. All authors contributed to the manuscript writing. All authors have read and approved the final version of the manuscript and agree with the order of presentation of the authors.

## Acknowledgements

This work was mainly supported by the Grant PID2020-120249RB-I00 funded by MCIN/AEI/10.13039/501100011033 and by the Andalusian Government (Junta de Andalucía, Plan Andaluz de Investigación, ref. P20_00124). IEC is supported by grant RYC2019-027287-I funded by MCIN/AEI/10.13039/501100011033/ and “ESFInvesting in your future”, and grants PID2022-137399OB-I00 and CNS2024-154835 funded by MCIN/AEI/10.13039/501100011033/ and “ERDF A way of making Europe”. JSM was supported by the National Agency for Research and Development (ANID)/Scholarship Program/DOCTORADO BECAS CHILE/2022–(Grant N°72220164). AT has received funding from the Junta de Andalucia, Spain, under the Post-doctoral Research Fellows (ref. POSTDOC_ 21_00745). BFG is supported by MCIN/AEI/10.13039/501100011033 and FSE+ (PID2022-137399OB-I00). MOR, JFO, ACP, and LSA are supported by the Spanish Ministry of Science, Innovation and Universities (FPU 22/02476, FPU 22/03052, FPU 21/0294, and FPU21/06192, respectively). EAB has received funding from the European Union’s Horizon 2020 research and innovation programme under the Marie Skłodowska-Curie grant agreement No (101064851). This work is part of a Ph.D. Thesis conducted in the Biomedicine Doctoral Studies of the University of Granada, Spain. The authors would like to thank the participants who took part in this study.

## Supplementary material for

### Supplementary Methods

The structural and diffusion MRI preprocessing procedures, as well as the quality control methods described below, are reproduced from an approach that we previously applied in an independent sample.^1^ To facilitate reproducibility and ensure that this Supplementary Methods section is self-contained, we include the same methodological description here. The details are provided below.

#### 1.1. T1-weighted MPRAGE MRI processing and measurements

##### 1.1.1. Image pre-processing

T1-weighted MPRAGE images were processed using FreeSurfer 7.4.1 (https://surfer.nmr.mgh.harvard.edu) on Neurodesk,^2^ with the following three steps for longitudinal processing.^3^ The pipeline is summarized in Supplementary Figure 1, and the script implementing the pipeline is publicly available on GitHub (https://github.com/Heart-Brain/Heart-Brain). First, in the cross-sectional processing, the two images (pre- and post-intervention) of each subject were independently processed using the “recon-all” pipeline, with the -3T flag (*recon-all -3T)*. Second, a within-subject template (*recon-all -base*) was created by averaging the processed images from the cross-sectional processing. Third, longitudinal processing used the outputs of the previous steps to obtain the final images (*recon-all -long*).

##### 1.1.2. Cortical thickness and volume extraction

Cortical thickness of brain regions was extracted from FreeSurfer’s outputs files (*lh.aparc.stats* and *rh.aparc.stats* files), while subcortical volumes for the left and right hemispheres, as well as the estimated intracranial volume, were obtained from the *aseg.stats* file. All files contain the Desikan-Killiany atlas parcellation.^4^

##### 1.1.3. Image quality check

Quality control (QC) was independently performed by two researchers (JSM, JOF) following the ENIGMA Consortium Cortical QC Protocol 2.0 (https://enigma.ini.usc.edu). Image quality of the FreeSurfer parcellation output was categorized as *pass*, *moderate* or *fail,* following ENIGMA protocol recommendations (https://enigma.ini.usc.edu/protocols/imaging-protocols/). See Results for a summary of QC results.

#### 1.2. Diffusion MRI processing and measurements

##### 1.2.1. Image pre-processing

DWI were processed on Neurodesk using command modules from MRtrix3 v3.04,^5^ FSL v6.0.7.16,^6^ and ANTs v2.6.0.^7^ The pipeline is summarized in Supplementary Figure 2, and the script implementing the pipeline is publicly available on GitHub (https://github.com/Heart-Brain/Heart-Brain). First, the diffusion-weighted gradient scheme (bvecs/bval files) was imported during the conversion of raw NifTi images to MRtrix3 format (.mif). The preprocessing steps included denoising (*dwidenoise* command), and removal of Gibbs ringing artifacts (*mrdegibbs* command). Motion and distortion correction with FSL’s eddy and topup tools within the *dwifslpreproc* script in MRtrix3, with phase encoding set to *-pe_dir j*, and including -eddy_options (“--repol^8^ --cnr_maps --slm=linear). Images acquired with a different phase encoding (n = 3) were excluded. Bias field correction was applied using the ANTs algorithm via the *dwibiascorrect* command in MRtrix3, and brain masking was done with SynthStrip (*mri_synthstrip*, v7.4.1).^9^ Diffusion tensors were computed using *dwi2tensor*, and mean diffusivity (MD) was calculated as the average of the 3 eigenvalues (λ₁, λ₂, λ₃) at each voxel. To avoid physically implausible values, negative eigenvalues were set to zero prior to computation, as described in previous studies.^10,11^

##### 1.2.2. Gray matter mean diffusivity processing

First, the b0 image (extracted from the preprocessed DWI) was registered to its respective T1 image (T1.mgz output from FreeSurfer) using the FSL’s *epi_reg* command.^12^ The resulting transformation matrix was then applied to the MD images using the *FLIRT* command. A spatial smoothing with a 6-mm full-width at half maximum (FWHM) Gaussian kernel was applied to the MD images using the *mrfilter* command.

To limit the analysis to gray matter voxels and minimize the potential impact of partial volume effects, individual masks for gray matter and cerebrospinal fluid (CSF) were generated. The gray matter mask was derived from each *aparc+aseg.mgz* file (FreeSurfer output) using the *5ttgen freesurfer* command in MRtrix3, which produced binary segmentations of cortical gray matter, sub-cortical gray matter, white matter, and CSF. The cortical and subcortical gray matter images were extracted, merged into a single mask, and then resampled to match the resolution and voxel grid of the corresponding FreeSurfer T1 image using *antsApplyTransforms* with nearest-neighbor interpolation (*--interpolation NearestNeighbor*) and an identity transformation *(-t identity*), ensuring no geometric transformation was applied.

As recommended,^13^ partial volume effects arising from CSF contamination must be addressed when analyzing GMMD. A CSF mask was created from the preprocessed DWI using multi-tissue constrained spherical deconvolution in MRtrix3, executed with the *dwi2fod msmt_csd* command for multi-shell diffusion data. This approach estimated the contribution of white matter-like, gray matter-like and free water CSF-like signals within each voxel. The three tissue compartment images were normalized using *mtnormalise*. The first volume of the white matter compartment was extracted using *mrconvert*, then summed with the gray matter and CSF signals using *mrcalc*. An image reflecting the proportion of CSF signal in each voxel was generated using *mrcalc*, and the b0-to-T1 transformation matrix was applied using the *FLIRT* command. Following prior research and recommendations,^14,15^ each subject’s CSF mask was binarized with a threshold of 0.5 (voxels with ≥50% CSF-like signal) with *mrcalc*. The final individual gray matter mask was generated by excluding any voxels overlapping with the CSF mask and was applied to the MD image to generate a filtered GMMD image, which contains MD values only in gray matter voxels (Supplementary Figure 2).

##### 1.2.3. Gray matter mean diffusivity extraction

Cortical and subcortical regions from the Desikan-Killiany parcellation atlas were generated for each subject using the *labelconvert* command in MRtrix3 applied to FreeSurfer’s *aparc+aseg.mgz* image. Individual masks for each region were generated with *mrcalc*. Finally, mean GMMD for each cortical and subcortical region was calculated by averaging GMMD values across all voxels within each region using *mrstats*.

##### 1.2.4. Image quality check

Two researchers (JSM, MTRP) independently assessed raw image quality using a 4-point scale:^16^ 1= “excellent”, 2 = “minor”, 3 = “moderate, and 4 = “severe”, based on motion and artifacts (e.g., spiking, ghosting, missing slices). Images rated as severe were excluded; those rated as moderate were included in the main analyses but excluded from sensitivity analyses. Additionally, automatic QC was performed using EDDY QC^17^ (*-eddyqc_all* in *dwifslpreproc,* MRtrix3). Extracted metrics included average absolute motion, average relative motion, outliers’ percentage, average contrast-to-noise-ratio (CNR), and signal-to-noise-ratio (SNR) in the b0. Images exceeding thresholds (absolute motion ≥2 mm, relative motion ≥0.5 mm, outliers ≥2%, SNR ≤20, CNR ≤1.5)^18^ in 2 or more metrics were excluded. See Results for a summary of QC results.

## Supplementary Results

### Quality control

Details on participant inclusion and exclusion based on brain image quality are summarized in Supplementary Table 3. After applying the ENIGMA protocol, most image parcellations were rated as *pass* (53.6%) or *moderate* (46.3%), with none rated as *fail*. Due to parcellation issues of ROIs in the *moderate* category, 8 participants’ images were excluded from the sensitivity analysis of the thickness/volume signature. For visual DWI QC, 46.9% participant’s images were rated as *excellent*, 48% as *minor*, 4.5% (8 images) as *moderate*, and 0.6% (1 image) as *severe*. Automatic QC did not identify low-quality images. Additionally, 2 images were obtained using an incorrect phase direction. Thus, 3 participants’ images were excluded from the main analysis, and an additional 15 participants’ images were excluded from the sensitivity analysis of the GMMD signature.

---

**Supplementary Figure 1.**
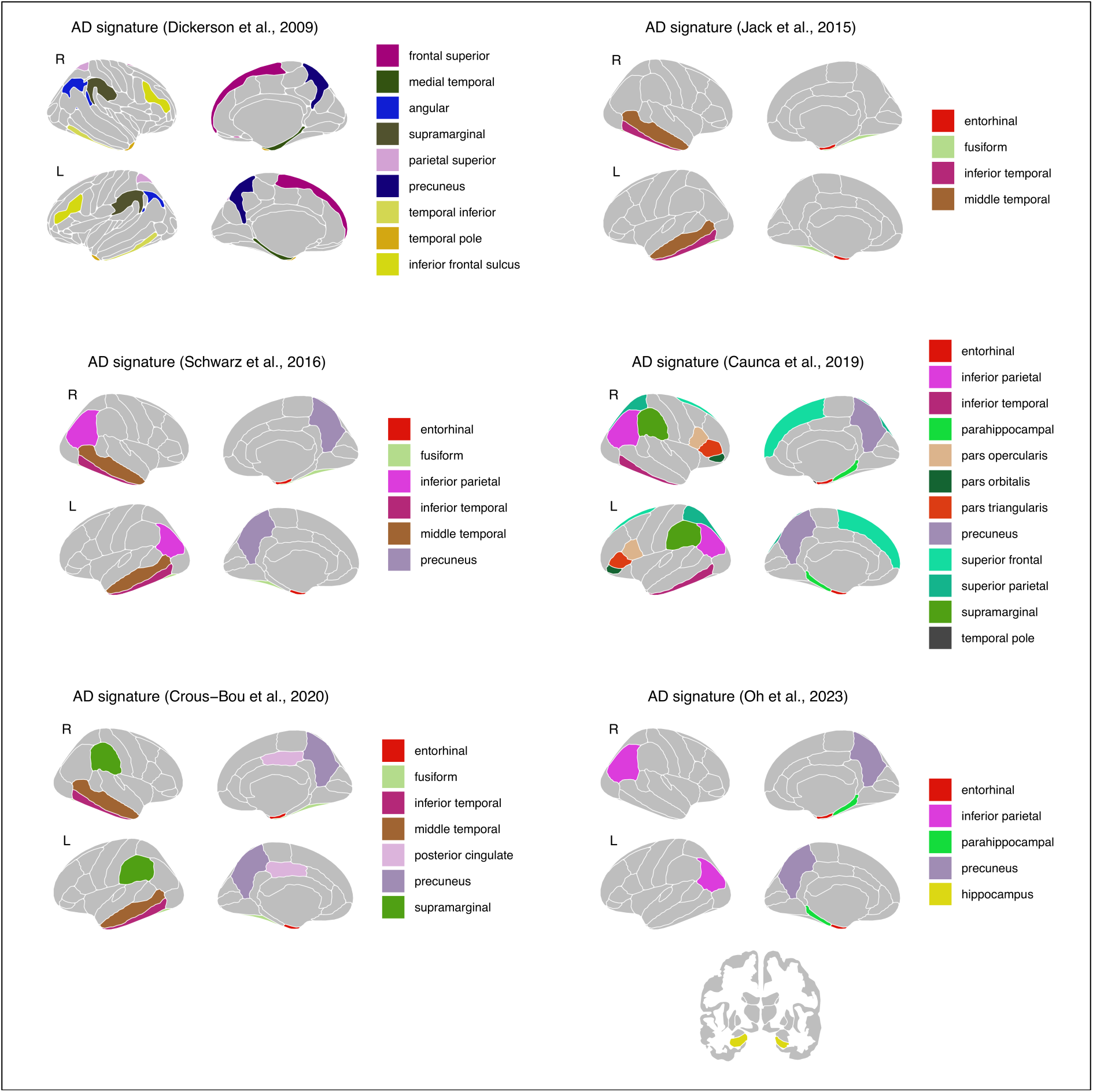
Brain regions used by the methodologies included to compute Alzheimer’s disease brain signatures. Most Alzheimer’s disease brain signatures were derived by averaging individual cortical thicknesses across bilateral regions of interest (ROIs) and then z-scored; however, Oh et al. (2023) utilized brain volumes. ROIs for the Jack et al. (2015), Schwarz et al. (2016), Caunca et al. (2019), Crous-Bou et al. (2020), and Oh et al. (2023) approaches were obtained from FreeSurfer’s outputs (‘lh.aparc.stats’ and ‘rh.aparc.stats’ files), which include the Desikan-Killiany atlas parcellation (Desikan et al., 2006). Additionally, hippocampal volumes for the Oh et al. (2023) approach were obtained from the ‘aseg.stats’ file (a FreeSurfer output), which also contains the Desikan-Killiany atlas parcellation. ROIs for the Dickerson et al. (2009) approach were obtained from FreeSurfer’s outputs (‘lh.aparc.a2009s.stats’ and ‘rh.aparc.a2009s.stats’ files), which include the Destrieux atlas parcellation (Destrieux et al., 2009). Abbreviation: AD, Alzheimer’s disease.

**Supplementary Figure 2.**
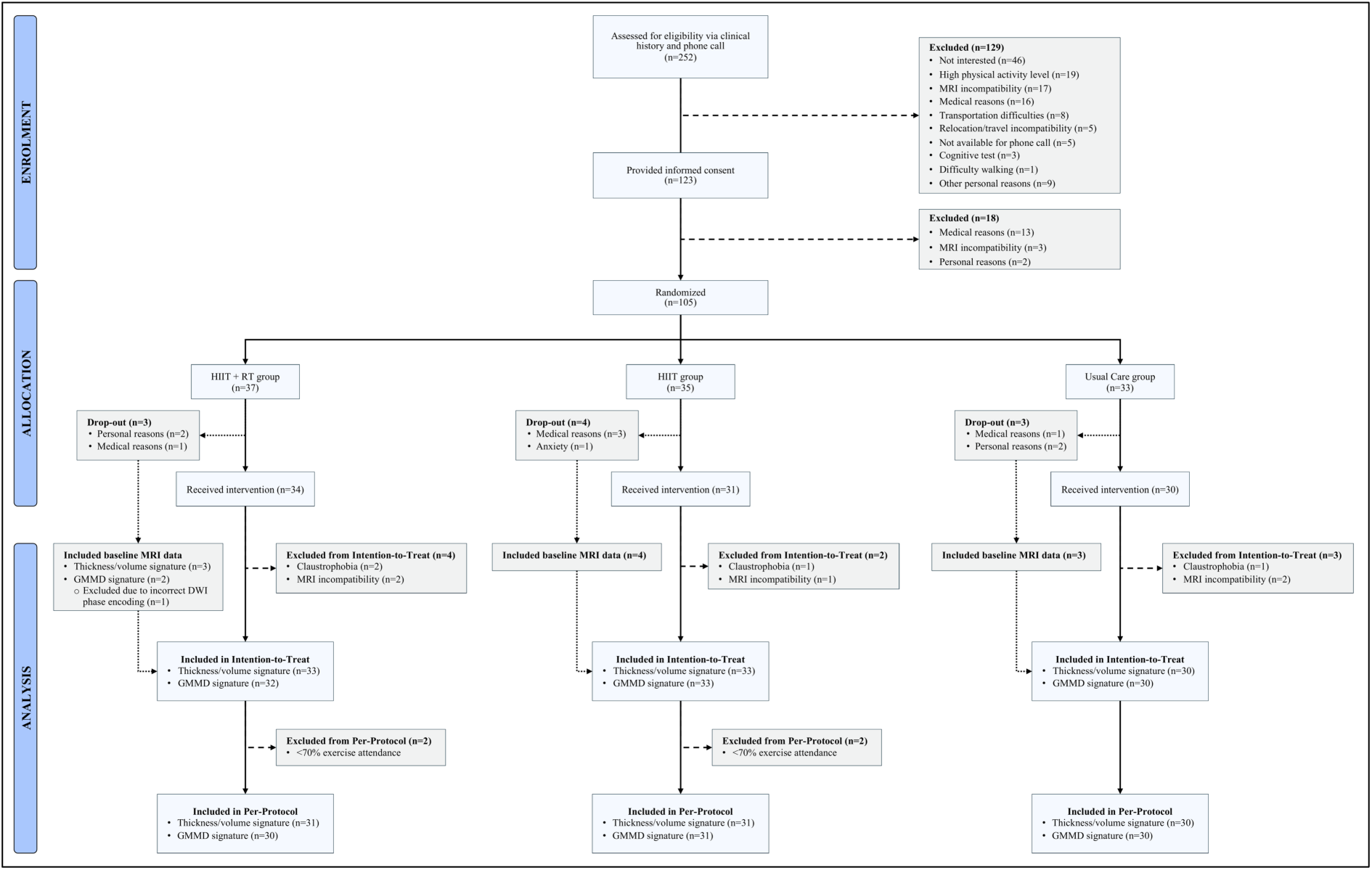
Consolidated Standards of Reporting Trials (CONSORT) flow diagram. DWI, diffusion-weighted images; GMMD, gray matter mean diffusivity; HIIT, high-intensity interval training; MRI, magnetic resonance imaging; RT, resistance training.

**Supplementary Table 1.** Consolidated Standards of Reporting Trials (CONSORT) checklist.

| Section/topic | No | CONSORT 2025 checklist item description | Reported on page no. |
| --- | --- | --- | --- |
| **Title and abstract** |  |  |  |
| Title and structured abstract | 1a | Identification as a randomised trial | 1 |
|  | 1b | Structured summary of the trial design, methods, results, and conclusions | 2 |
| **Open science** |  |  |  |
| Trial registration | 2 | Name of trial registry, identifying number (with URL) and date of registration | 4 |
| Protocol and statistical analysis plan | 3 | Where the trial protocol and statistical analysis plan can be accessed | 4 |
| Data sharing | 4 | Where and how the individual de-identified participant data (including data dictionary), statistical code and any other materials can be accessed | 4 |
| Funding and conflicts of interest | 5a | Sources of funding and other support (eg, supply of drugs), and role of funders in the design, conduct, analysis and reporting of the trial | 12 |
|  | 5b | Financial and other conflicts of interest of the manuscript authors | 12 |
| **Introduction** |  |  |  |
| Background and rationale | 6 | Scientific background and rationale | 3 |
| Objectives | 7 | Specific objectives related to benefits and harms | 4 |
| **Methods** |  |  |  |
| Patient and public involvement | 8 | Details of patient or public involvement in the design, conduct and reporting of the trial | Ref 20 |
| Trial design | 9 | Description of trial design including type of trial (eg, parallel group, crossover), allocation ratio, and framework (eg, superiority, equivalence, non-inferiority, exploratory) | 4 |
| Changes to trial protocol | 10 | Important changes to the trial after it commenced including any outcomes or analyses that were not prespecified, with reason | 4 |
| Trial setting | 11 | Settings (eg, community, hospital) and locations (eg, countries, sites) where the trial was conducted | 4,5 |
| Eligibility criteria | 12a | Eligibility criteria for participants | 4 |
|  | 12b | If applicable, eligibility criteria for sites and for individuals delivering the interventions (eg, surgeons, physiotherapists) | Ref 19, Ref 24 |
| Intervention and comparator | 13 | Intervention and comparator with sufficient details to allow replication. If relevant, where additional materials describing the intervention and comparator (eg, intervention manual) can be accessed | 5, Ref 24 |
| Outcomes | 14 | Prespecified primary and secondary outcomes, including the specific measurement variable (eg, systolic blood pressure), analysis metric (eg, change from baseline, final value, time to event), method of aggregation (eg, median, proportion), and time point for each outcome | 4,5,6 |
| Harms | 15 | How harms were defined and assessed (eg, systematically, non-systematically) | 7, Ref 20 |
| Sample size | 16a | How sample size was determined, including all assumptions supporting the sample size calculation | 4 |
|  | 16b | Explanation of any interim analyses and stopping guidelines | Statistical Analysis Plan |
| Randomisation: |  |  |  |
| Sequence generation | 17a | Who generated the random allocation sequence and the method used | 4, Ref 19 |
|  | 17b | Type of randomisation and details of any restriction (eg, stratification, blocking and block size) | 4, Ref 19 |
| Allocation concealment mechanism | 18 | Mechanism used to implement the random allocation sequence (eg, central computer/telephone; sequentially numbered, opaque, sealed containers), describing any steps to conceal the sequence until interventions were assigned | 4, Ref 19 |
| Implementation | 19 | Whether the personnel who enrolled and those who assigned participants to the interventions had access to the random allocation sequence | 4, Ref 19 |
| Blinding | 20a | Who was blinded after assignment to interventions (eg, participants, care providers, outcome assessors, data analysts) | 4, Ref 19 |
|  | 20b | If blinded, how blinding was achieved and description of the similarity of interventions | Ref 19 |
| Statistical methods | 21a | Statistical methods used to compare groups for primary and secondary outcomes, including harms | 6,7 |
|  | 21b | Definition of who is included in each analysis (eg, all randomised participants), and in which group | 6 |
|  | 21c | How missing data were handled in the analysis | 6,7 |
|  | 21d | Methods for any additional analyses (eg, subgroup and sensitivity analyses), distinguishing prespecified from post hoc | 7 |
| **Results** |  |  |  |
| Participant flow, including flow diagram | 22a | For each group, the numbers of participants who were randomly assigned, received intended intervention, and were analysed for the primary outcome | Supplementary Figure 2 |
|  | 22b | For each group, losses and exclusions after randomisation, together with reasons | Supplementary Figure 2, Supplementary Table 3 |
| Recruitment | 23a | Dates defining the periods of recruitment and follow-up for outcomes of benefits and harms | 4 |
|  | 23b | If relevant, why the trial ended or was stopped | NA |
| Intervention and comparator delivery | 24a | Intervention and comparator as they were actually administered (eg, where appropriate, who delivered the intervention/comparator, how participants adhered, whether they were delivered as intended (fidelity)) | 5,7, Ref 20 |
|  | 24b | Concomitant care received during the trial for each group | Ref 20 |
| Baseline data | 25 | A table showing baseline demographic and clinical characteristics for each group | Table 1 |
| Numbers analysed, outcomes and estimation | 26 | For each primary and secondary outcome, by group:<br>● the number of participants included in the analysis<br>● the number of participants with available data at the outcome time point<br>● result for each group, and the estimated effect size and its precision (such as 95% confidence interval)<br>● for binary outcomes, presentation of both absolute and relative effect size | Supplementary Figure 2, Supplementary Table 3, 8, Figure 2 |
| Harms | 27 | All harms or unintended events in each group | 7, Ref 20 |
| Ancillary analyses | 28 | Any other analyses performed, including subgroup and sensitivity analyses, distinguishing pre-specified from post hoc | 8, Figures 3-4, Supplementary Tables 4-7 |
| **Discussion** |  |  |  |
| Interpretation | 29 | Interpretation consistent with results, balancing benefits and harms, and considering other relevant evidence | 9, 10, 11 |
| Limitations | 30 | Trial limitations, addressing sources of potential bias, imprecision, generalisability, and, if relevant, multiplicity of analyses | 11 |

**Supplementary Table 2.** Cognitive outcomes and their corresponding cognitive tests.

| Domain | Cognitive test | Outcomes |
| --- | --- | --- |
| General cognition | MoCA | MoCA total score |
| Episodic memory | MoCA | Delayed recall |
|  | Picture Sequence Memory Test | Raw score |
| Executive function /<br>Attentional control | Dimensional Change Card Sort Task | Computed score |
|  | Flanker | Computed score |
|  | Trail Making Test | TMT-B |
| Processing speed | Digit Symbol Substitution Test | Total correct raw score |
|  | Trail Making Test | Time (part A) |
| Working memory | List Sorting Working Memory Test | Raw score |
|  | Spatial Working Memory Task | 4-item accuracy (high-load) |

Abbreviations: MoCA, Montreal Cognitive Assessment.

**Supplementary Table 3.** Inclusion and exclusion of participants’ brain images considering image quality for main and sensitivity <u>analyses by Alzheimer’s disease brain signature.</u>.

|  | HIIT+RT |  | HITT |  | UC |  | Total images |
| --- | --- | --- | --- | --- | --- | --- | --- |
|  | Pre | Post | Pre | Post | Pre | Post |  |
| *For thickness/volume signature analyses* |  |  |  |  |  |  |  |
| All T1 images | 33 | 29 | 33 | 29 | 30 | 25 | 179 |
| **Included for main analysis** | **33** | **29** | **33** | **29** | **30** | **25** | **179** |
| Moderate QC rating with issues in parcellation of ROIs | 2 | 1 | 1 | 1 | 2 | 1 | 8 |
| **Included for sensitive analysis** | **31** | **28** | **32** | **28** | **28** | **24** | **171** |
| *For GMMD signature analyses* |  |  |  |  |  |  |  |
| All DWI | 33 | 29 | 33 | 29 | 30 | 25 | 179 |
| Incorrect phase direction | 2 | 0 | 0 | 0 | 0 | 0 | 2 |
| Visual DWI QC rated as severe | 0 | 0 | 0 | 1 | 0 | 0 | 1 |
| Automatic QC $\geq 2$ exceeded thresholds | 0 | 0 | 0 | 0 | 0 | 0 | 0 |
| **Included for main analysis** | **31** | **29** | **33** | **28** | **30** | **25** | **176** |
| Parcellation issues of ROIs in the FreeSurfer output | 2 | 1 | 1 | 1 | 2 | 1 | 8 |
| Visual DWI QC rated as moderate | 1 | 0 | 3<sup>a</sup> | 3 | 1 | 0 | 8<sup>a</sup> |
| **Included for sensitive analysis** | **28** | **28** | **30** | **24** | **27** | **24** | **161** |

Abbreviations: DWI, diffusion-weighted images; GMMD, gray matter mean diffusivity; HIIT, high-intensity interval training; QC, quality control; ROIs, regions of interest; RT, resistance training; UC, usual care group.
<sup>a</sup> One image corresponds to a participant image with parcellation issues in the FreeSurfer output.

**Supplementary Table 4.** Estimated marginal means in Alzheimer’s disease brain signatures, excluding patients with <70% attendance (per-protocol analysis).

| Outcome | Within-group differences (estimate [95% CI]) |  |  | Between-groups differences (estimate [95% CI]) |  |  |  |  |  |
| --- | --- | --- | --- | --- | --- | --- | --- | --- | --- |
|  | HIIT+RT | HIIT | UC | HIIT+RT vs UC | p | HIIT vs UC | p | HIIT+RT vs HIIT | p |
| Thickness/volume signature | 0.16 [-0.03; 0.35] | -0.03 [-0.22; 0.16] | 0.06 [-0.14; 0.25] | 0.1 [-0.1; 0.3] | 0.32 | -0.09 [-0.29; 0.11] | 0.4 | 0.19 [-0.01; 0.39] | 0.06 |
| GMMD signature | 0.05 [-0.2; 0.3] | 0.09 [-0.16; 0.34] | -0.03 [-0.29; 0.23] | 0.08 [-0.18; 0.34] | 0.56 | 0.12 [-0.15; 0.38] | 0.39 | -0.04 [-0.3; 0.23] | 0.78 |

Data are presented as mean change [95% confidence interval]. Abbreviations: GMMD, gray matter mean diffusivity; HIIT, high-intensity interval training; RT, resistance training; UC, usual care group.

**Supplementary Table 5.** Estimated marginal means in Alzheimer’s disease brain signatures, excluding low-quality images (sensitivity analysis).

| Outcome | Within-group differences (estimate [95% CI]) |  |  | Between-groups differences (estimate [95% CI]) |  |  |  |  |  |
| --- | --- | --- | --- | --- | --- | --- | --- | --- | --- |
|  | HIIT+RT | HIIT | UC | HIIT+RT vs UC | p | HIIT vs UC | p | HIIT+RT vs HIIT | p |
| Thickness/volume signature | 0.21 [0.02; 0.39] | -0.07 [-0.25; 0.12] | 0.03 [-0.17; 0.23] | 0.17 [-0.03; 0.38] | 0.1 | -0.1 [-0.3; 0.1] | 0.34 | 0.27 [0.07; 0.47] | **0.01** |
| GMMD signature | 0.09 [-0.17; 0.35] | 0.13 [-0.15; 0.4] | -0.05 [-0.32; 0.22] | 0.14 [-0.13; 0.41] | 0.32 | 0.18 [-0.1; 0.46] | 0.22 | -0.04 [-0.31; 0.24] | 0.78 |

Data are presented as mean change [95% confidence interval]. Significant results shown in bold ( $p < 0.05$ ). Abbreviations: GMMD, gray matter mean diffusivity; HIIT, high-intensity interval training; RT, resistance training; UC, usual care group.

**Supplementary Table 6.** Estimated marginal means in Alzheimer’s disease brain signatures, excluding participants that only had pre-intervention images (sensitivity analysis).

| Outcome | Within-group differences (estimate [95% CI]) |  |  | Between-groups differences (estimate [95% CI]) |  |  |  |  |  |
| --- | --- | --- | --- | --- | --- | --- | --- | --- | --- |
|  | HIIT+RT | HIIT | UC | HIIT+RT vs UC | p | HIIT vs UC | p | HIIT+RT vs HIIT | p |
| Thickness/volume signature | 0.13 [-0.02; 0.28] | -0.12 [-0.26; 0.03] | -0.02 [-0.17; 0.14] | 0.15 [-0.06; 0.36] | 0.18 | -0.1 [-0.31; 0.11] | 0.34 | 0.25 [0.05; 0.45] | **0.02** |
| GMMD signature | 0.04 [-0.15; 0.22] | 0.06 [-0.12; 0.24] | -0.04 [-0.24; 0.15] | 0.08 [-0.18; 0.34] | 0.54 | 0.1 [-0.16; 0.36] | 0.44 | -0.02 [-0.27; 0.23] | 0.86 |

Data are presented as mean change [95% confidence interval]. Significant results shown in bold ( $p < 0.05$ ). Abbreviations: GMMD, gray matter mean diffusivity; HIIT, high-intensity interval training; RT, resistance training; UC, usual care group.

**Supplementary Table 7.**
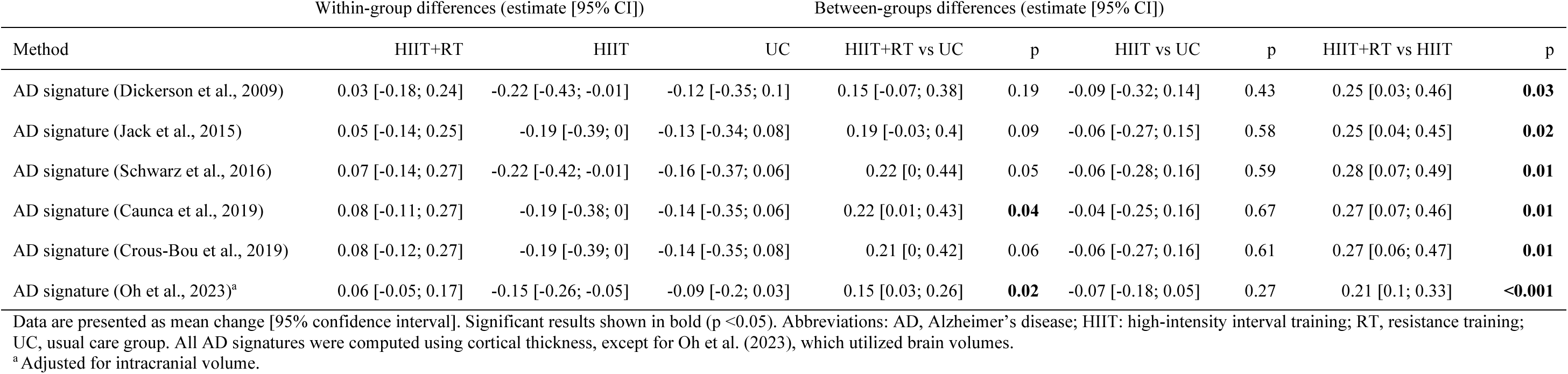
Estimated marginal means in Alzheimer’s disease signature cortical thickness or volume using additional methodologies.

| Method | Within-group differences (estimate [95% CI]) |  |  | Between-groups differences (estimate [95% CI]) |  |  |  |  |  |
| --- | --- | --- | --- | --- | --- | --- | --- | --- | --- |
|  | HIIT+RT | HIIT | UC | HIIT+RT vs UC | p | HIIT vs UC | p | HIIT+RT vs HIIT | p |
| AD signature (Dickerson et al., 2009) | 0.03 [-0.18; 0.24] | -0.22 [-0.43; -0.01] | -0.12 [-0.35; 0.1] | 0.15 [-0.07; 0.38] | 0.19 | -0.09 [-0.32; 0.14] | 0.43 | 0.25 [0.03; 0.46] | **0.03** |
| AD signature (Jack et al., 2015) | 0.05 [-0.14; 0.25] | -0.19 [-0.39; 0] | -0.13 [-0.34; 0.08] | 0.19 [-0.03; 0.4] | 0.09 | -0.06 [-0.27; 0.15] | 0.58 | 0.25 [0.04; 0.45] | **0.02** |
| AD signature (Schwarz et al., 2016) | 0.07 [-0.14; 0.27] | -0.22 [-0.42; -0.01] | -0.16 [-0.37; 0.06] | 0.22 [0; 0.44] | 0.05 | -0.06 [-0.28; 0.16] | 0.59 | 0.28 [0.07; 0.49] | **0.01** |
| AD signature (Caunca et al., 2019) | 0.08 [-0.11; 0.27] | -0.19 [-0.38; 0] | -0.14 [-0.35; 0.06] | 0.22 [0.01; 0.43] | **0.04** | -0.04 [-0.25; 0.16] | 0.67 | 0.27 [0.07; 0.46] | **0.01** |
| AD signature (Crous-Bou et al., 2019) | 0.08 [-0.12; 0.27] | -0.19 [-0.39; 0] | -0.14 [-0.35; 0.08] | 0.21 [0; 0.42] | 0.06 | -0.06 [-0.27; 0.16] | 0.61 | 0.27 [0.06; 0.47] | **0.01** |
| AD signature (Oh et al., 2023)<sup>a</sup> | 0.06 [-0.05; 0.17] | -0.15 [-0.26; -0.05] | -0.09 [-0.2; 0.03] | 0.15 [0.03; 0.26] | **0.02** | -0.07 [-0.18; 0.05] | 0.27 | 0.21 [0.1; 0.33] | **<0.001** |

Data are presented as mean change [95% confidence interval]. Significant results shown in bold ( $p < 0.05$ ). Abbreviations: AD, Alzheimer's disease; HIIT: high-intensity interval training; RT, resistance training; UC, usual care group. All AD signatures were computed using cortical thickness, except for Oh et al. (2023), which utilized brain volumes.
<sup>a</sup> Adjusted for intracranial volume.

